# Heart Rate Variability as a Biomarker of Burnout in Healthcare Workers: A Predictive Model Integrating Psychosocial and Occupational Factors

**DOI:** 10.1101/2025.09.06.25335221

**Authors:** Alberto Rubio-López, Alejandro Rubio-Navas, Teresa Sierra-Puerta, Rodrigo García-Carmona

## Abstract

**Background:** Burnout is a significant concern among healthcare professionals, particularly in high-stress environments such as intensive care units (ICUs). While prior research has linked burnout to self-reported stress and psychological distress, objective physiological markers like heart rate variability (HRV) may offer a more reliable assessment of occupational stress and burnout risk. Our previous pilot study suggested an association between HRV and stress; however, it did not incorporate standardized burnout assessments. This study aims to bridge that gap by examining the relationship between HRV, self-reported stress, and validated burnout scales. Additionally, it seeks to identify key predictors of burnout and develop a predictive model for early risk detection.

**Methods:** This cross-sectional observational study included 57 nurses and nursing assistants working in ICUs and general hospital wards. Participants completed validated burnout assessments, including the Cuestionario para la Evaluación del Síndrome de Quemarse por el Trabajo (CESQT; Spanish Burnout Inventory), the Maslach Burnout Inventory (MBI), the Professional Quality of Life Scale (ProQOL), and the State-Trait Anxiety Inventory (STAI). HRV parameters were recorded using a Biosignals Plux system for 10 minutes at rest before the start of the work shift and analyzed with the OpenSignals software. Extracted HRV metrics included the root mean square of successive differences (rMSSD), low-frequency to high-frequency ratio (LF/HF), Standard Deviation 1 and 2 Ratio (SD1/SD2 ratio), and Poincaré area. Statistical analyses involved descriptive statistics, correlation analysis, and group comparisons to examine differences in burnout across workplace conditions, shift types, and shift durations. A logistic regression model with 10-fold cross-validation was developed to predict burnout risk, integrating HRV parameters, psychological distress, and occupational factors.

**Results:** HRV parameters were significantly associated with self-reported stress and burnout indicators, reinforcing their potential role as objective biomarkers of occupational stress. Night shift workers and those with extended work hours exhibited higher burnout levels and greater autonomic dysregulation. The predictive model demonstrated strong accuracy in identifying individuals at risk of burnout. The model integrating HRV parameters, psychological distress, and occupational factors (Model 2) achieved an AUC-ROC of 0.832 (95% CI: 0.735– 0.929) and an accuracy of 79.1%, outperforming the model based solely on demographic and psychometric data (Model 1, AUC-ROC = 0.791, 95% CI: 0.685–0.897, accuracy = 76.3%). HRV and psychological stress emerged as key contributing factors.

**Conclusion:** These findings highlight HRV as a promising tool for the objective assessment of burnout risk in healthcare professionals. The predictive model developed provides a framework for early identification of high-risk individuals, enabling targeted interventions to improve well-being and staff retention in healthcare settings. Future research should validate these findings in larger cohorts and assess the long-term applicability of HRV-based monitoring systems in occupational health programs.

## INTRODUCTION

Burnout syndrome is an increasingly recognized occupational health concern, particularly among healthcare professionals exposed to high workloads, emotional strain, and persistent stress conditions [1]. Characterized by emotional exhaustion, depersonalization, and diminished personal accomplishment, burnout is associated with increased absenteeism, staff turnover, and reduced quality of patient care. Given its significant impact on individual well-being and healthcare system performance, the early identification of objective indicators of burnout has become a priority in occupational health research [2].

Traditional burnout assessments rely primarily on self-reported questionnaires, such as the Maslach Burnout Inventory (MBI) [3] and the *Cuestionario para la Evaluación del Síndrome de Quemarse por el Trabajo* (CESQT Spanish Burnout Inventory) [4]. Although these tools are well validated, self-report measures are inherently subject to biases, including self-perception distortions, response tendencies, and social desirability effects. These limitations underscore the need for objective physiological indicators that can provide more accurate and real-time assessments of occupational stress and burnout risk.

Heart rate variability (HRV) has emerged as a promising physiological marker for evaluating autonomic nervous system (ANS) activity and stress adaptation capacity. HRV reflects the dynamic balance between sympathetic and parasympathetic branches of the ANS and serves as a non-invasive index of autonomic flexibility [5]. Reduced HRV has been consistently associated with chronic stress, anxiety disorders, and occupational burnout, suggesting its potential as a biomarker for identifying individuals at risk. Specific HRV parameters, such as the Standard Deviation 1 and 2 (SD1/SD2) ratio, root mean square of successive differences (rMSSD), and low-frequency/high-frequency (LF/HF) ratio, have shown associations with work-related stress, fatigue, and emotional exhaustion [6].

A prior pilot study conducted among ICU nurses by our team found significant correlations between HRV indices and subjective stress levels [7], suggesting that HRV may function as a physiological marker of occupational distress. However, that study did not incorporate standardized burnout instruments, limiting its interpretability. Moreover, despite supportive findings, HRV has not yet been widely implemented as a screening tool in occupational health contexts.

In addition to individual physiological responses, organizational and shift-related factors play a critical role in burnout development. Night shifts and extended work hours have been linked to increased psychological distress and reduced HRV, likely due to circadian disruption and sustained physical and cognitive demand [8]. Additionally, burnout prevalence varies across professional roles, with ICU nurses experiencing higher stress levels than general ward nurses due to frequent exposure to critically ill patients, complex decision-making, and heavier workloads. Similar trends have been observed among nursing assistants, yet they remain underrepresented in burnout research. Furthermore, professionals expressing an intention to leave the profession tend to show lower HRV and higher burnout scores, reinforcing the association between job dissatisfaction and physiological dysregulation [9].

### Study Objectives

Building upon the growing evidence linking HRV and burnout, this study aimed to develop and evaluate a predictive model that integrates physiological, psychological, and occupational factors to support the early identification of healthcare professionals at risk of burnout [10].

To overcome the limitations of self-reported assessments, we designed two logistic regression models: 1) Model 1: incorporating demographic and psychometric data (e.g., anxiety levels, burnout scores); and 2) Model 2: integrating HRV parameters, shift type, and additional occupational variables.

Both models were validated using a 10-fold cross-validation to ensure generalizability and robustness. By combining biometric, behavioral, and work-related data, this study provides a comprehensive and objective approach to burnout risk detection in high-demand clinical environments.

## METHODS

### Study Design and Participants

This cross-sectional observational study was conducted with a sample of 57 nurses and nursing assistants working in intensive care units (ICUs) and general hospital wards. The mean age was 35.77 years (range: 24–63), and 68.4% were female. Participants were required to have at least six months of professional experience and be employed full-time at the time of data collection. Exclusion criteria included a history of cardiovascular diseases that could affect heart rate variability (HRV) or the presence of severe psychiatric disorders that might interfere with stress and burnout assessments.

No formal sample size calculation was performed prior to data collection, as the study was designed as an exploratory observational analysis. The sample size was based on the availability of eligible participants during the recruitment period and was considered adequate for preliminary correlation and predictive modeling. However, results should be interpreted with caution and validated in larger, independent cohorts.

### Ethical Considerations and Recruitment

The study was approved by the Research Ethics Committee of HM University Hospital (Code: 18.12.1339.GHM) and conducted in accordance with the Declaration of Helsinki. Data collection took place between September 1, 2024, and December 27, 2024. All participants provided written informed consent, and their data were fully anonymized to ensure confidentiality.

### Variables and Measurements

A comprehensive set of sociodemographic, occupational, psychological, and biometric variables was collected to examine potential predictors of burnout. Participants provided information on age, sex, job role (nurse or assistant nurse), work unit (ICU or general ward), shift type (day or night shift), and shift duration (8-hour or 12-hour shifts). Additional variables included self-reported health conditions (e.g., hypertension, diabetes, dyslipidemia, depression, anxiety) and lifestyle factors such as alcohol and tobacco use, recreational drug use, and physical activity. Indicators of work-related stress included perceived stress, suicidal ideation, and intention to leave the profession.

HRV parameters were recorded using a Biosignals Plux system (PLUX Wireless Biosignals S.A., Lisbon, Portugal). Measurements were taken for 10 minutes at rest before the start of each participant’s work shift ensuring a standardized baseline assessment. Data collection was conducted in a quiet environment, with participants seated comfortably to minimize external influences on autonomic activity. Data processing and analysis were performed using the OpenSignals software (version 2023-10-23) to ensure standardized signal processing and artifact filtering.

The following HRV parameters were extracted:

- Root mean square of successive differences (rMSSD): A marker of parasympathetic nervous system activity and short-term HRV.
- Percentage of successive RR intervals differing by more than 50 ms (PNN50) and 20 ms (PNN20): Indicators of vagal tone and cardiac autonomic flexibility.
- Low-frequency to high-frequency ratio (LF/HF): Reflects autonomic balance between sympathetic and parasympathetic systems.
- SD1/SD2 ratio: Nonlinear HRV parameter representing autonomic adaptability and resilience.

Burnout and psychological stress were assessed using four validated self-report instruments (see Supplementary Appendix A for full questionnaires)

- Cuestionario para la Evaluación del Síndrome de Quemarse por el Trabajo (CESQT): This tool measures four dimensions of burnout: work engagement (Ilusión por el Trabajo, IT), psychological exhaustion (Desgaste Psíquico, DP), indolence (Indolencia, IN), and guilt (Culpa, CU). This instrument was chosen as the primary burnout measure due to its inclusion of dimensions particularly relevant to healthcare professionals [11].
- Maslach Burnout Inventory (MBI): Assesses emotional exhaustion (EE), depersonalization (DP), and professional inefficacy (PI) [12].
- Professional Quality of Life Scale (ProQOL): Measures compassion satisfaction (CS), secondary traumatic stress (STS), and burnout (BO) [13].
- State-Trait Anxiety Inventory (STAI): Includes two subscales assessing state anxiety (STAI-AE) and trait anxiety (STAI-AR), providing insight into acute and chronic stress levels [14].

All psychometric instruments were administered prior to the start of the work shift, concurrent with HRV data collection, to ensure that responses reflected baseline psychological states rather than shift-related stressors.

### Statistical Analysis

Statistical analyses were performed in four sequential steps using IBM SPSS Statistics v. 25. First, descriptive analyses were conducted to summarize the data, including means and standard deviations for continuous variables and frequencies and percentages for categorical variables. Kolmogorov-Smirnov and Shapiro-Wilk tests confirmed that HRV parameters and burnout scores were non-normally distributed, leading to the use of non-parametric statistical methods. Second, Spearman’s rank-order correlations were used to evaluate associations between HRV parameters, stress and burnout scores (STAI, CESQT, MBI, ProQOL), and self-reported stress indicators. This step aimed to determine whether HRV could serve as an objective biomarker for burnout. Third, group comparisons were performed using non-parametric tests. Mann-Whitney U tests were conducted to compare HRV and burnout scores between day and night shift workers and between ICU and general ward nurses. Kruskal-Wallis tests were used to assess burnout differences based on shift duration (8-hour vs. 12-hour shifts).

Finally, multiple logistic regression analysis was applied to identify key predictors of burnout, with the CESQT Burnout Index as the dependent variable. HRV parameters, perceived stress, and occupational factors were included as independent variables. Two models were tested: 1) Model 1: Included demographic and psychometric predictors (e.g., age, gender, anxiety levels, burnout scores); and 2) Model 2: Incorporated HRV parameters, shift type, and additional occupational factors (e.g., self-perceived stress, work conditions).

To ensure model robustness, a 10-fold cross-validation procedure was implemented, and its performance was evaluated based on accuracy, sensitivity, specificity, and the area under the receiver operating characteristic curve (AUC-ROC).

## RESULTS

The study included 57 nurses and nursing assistants working in ICUs and general hospital wards, with a mean age of 35.77 years (range: 24–63), 68.4% female and an average professional experience of 4.67 years (56.05 months). Regarding mental health, 36.8% of participants reported anxiety, while 10.5% were diagnosed with depression. Suicidal ideation was present in 28.1%, highlighting severe psychological distress. Medication use was also notable, with 26.3% of participants using anxiolytics and 19.3% using antidepressants.

In terms of lifestyle factors, 21.1% of participants were smokers, while 78.9% consumed caffeine regularly. Only 31.6% engaged in regular physical activity, a factor known to influence stress regulation and burnout risk. Table 1 summarizes the demographic, occupational, and health-related characteristics of the sample, including subgroup comparisons between ICU and general ward staff.

**Table 1.** Demographic, occupational, and health characteristics of the study participants, including job roles, work unit, shift type, and mental health and lifestyle factors relevant to stress and burnout analysis. Values are presented as percentages (%) for the total sample and for ICU and general ward subgroups.

| Variable | Total (%) | ICU (%) | General Ward (%) |
| --- | --- | --- | --- |
| **Job Role** |  |  |  |
| Nurse (DUE) | 54.4 |  |  |
| Assistant Nurse (TCAE) | 45.6 |  |  |
| **Work Unit** |  |  |  |
| ICU | 52.6 |  |  |
| General Ward | 47.4 |  |  |
| **Work Shift** |  |  |  |
| Day shift | 56.1 |  |  |
| Night shift | 43.9 |  |  |
| **Shift Duration** |  |  |  |
| 8 hours | 49.1 |  |  |
| 12 hours | 50.9 |  |  |
| **Mental Health Conditions** |  |  |  |
| Anxiety | 36.8 | 43.33 | 29.63 |
| Depression | 10.5 | 6.67 | 14.81 |
| Suicidal Ideation | 28.1 | 30.0 | 25.93 |
| **Medication Use** |  |  |  |
| Anxiolytics | 26.3 | 16.67 | 37.04 |
| Antidepressants | 19.3 | 26.67 | 11.11 |
| **Lifestyle Factors** |  |  |  |
| Regular Physical Activity | 31.6 | 13.33 | 18.52 |
| Smokers | 21.1 | 66.67 | 74.07 |
| Caffeine Consumption | 78.9 | 13.33 | 11.11 |

### Burnout, Anxiety, and HRV Metrics

Burnout was assessed using both global indices and individual subscales from the CESQT, allowing for a multidimensional analysis of emotional engagement, exhaustion, indolence, and guilt. Participants exhibited moderate-to-high levels of burnout, with a mean CESQT Burnout Index of 35.42 ± 5.06. Within the Maslach Burnout Inventory (MBI), the Emotional Exhaustion subscale (23.02 ± 8.72) had the highest scores, followed by Depersonalization (7.7 ± 4.0) and Personal Accomplishment (25.18 ± 5.08). Anxiety symptoms were also prevalent, with state anxiety (STAI-AE) averaging 42.86 ± 10.24 and trait anxiety (STAI-AR) at 32.21 ± 8.88. Notably, 59.6% of participants expressed an intention to leave the profession, underscoring occupational dissatisfaction. HRV analysis revealed significant variability among participants. The mean RMSSD was 229.77 ± 104.86 ms, with a PNN50 of 59.14 ± 28.67% and an LF/HF ratio of 0.89 ± 1.24. The SD1/SD2 ratio, a measure of autonomic adaptability, averaged 0.83 ± 0.23. Table 2 presents a summary of HRV and psychometric assessments, including CESQT subscale scores and comparisons between ICU and general ward staff.

**Table 2.** Summary of heart rate variability (HRV) parameters and psychometric assessments related to stress, burnout, and anxiety. The table presents mean values and standard deviations (Mean ± SD) for the total sample, as well as for ICU and general ward subgroups. Included measures cover autonomic regulation (HRV), burnout dimensions (CESQT, MBI), and anxiety symptoms (STAI).

| Variable | Total<br>(Mean ± SD) | ICU<br>(Mean ± SD) | General Ward<br>(Mean ± SD) |
| --- | --- | --- | --- |
| RMSSD (ms) | 229.77 ± 104.86 | 210.33 ± 117.69 | 251.37 ± 85.52 |
| PNN50 (%) | 59.14 ± 28.67 | 48.87 ± 28.91 | 70.56 ± 24.1 |
| LF/HF Ratio | 0.89 ± 1.24 | 1.23 ± 1.62 | 0.52 ± 0.34 |
| SD1/SD2 Ratio | 0.83 ± 0.23 | 0.77 ± 0.26 | 0.89 ± 0.16 |
| CESQT – Burnout Index | 35.42 ± 5.06 | 36.43 ± 5.34 | 34.3 ± 4.57 |
| CESQT – Work Engagement (IT) | 9.82 ± 1.39 | 9.8 ± 1.49 | 9.85 ± 1.29 |
| CESQT – Psychological Exhaustion (DP) | 13.93 ± 2.45 | 14.17 ± 2.53 | 13.67 ± 2.37 |
| CESQT – Indolence (IN) | 11.32 ± 3.78 | 12.07 ± 4.08 | 10.48 ± 3.3 |
| CESQT – Guilt (CU) | 1.86 ± 1.43 | 1.67 ± 0.76 | 2.07 ± 1.92 |
| MBI – Emotional Exhaustion | 23.02 ± 8.73 | 25.33 ± 8.47 | 20.44 ± 8.43 |
| MBI – Depersonalization | 7.7 ± 4.0 | 8.63 ± 3.92 | 6.67 ± 3.9 |
| MBI – Personal Accomplishment | 25.18 ± 5.08 | 24.07 ± 4.99 | 26.41 ± 4.99 |
| STAI – State Anxiety | 42.86 ± 10.24 | 45.33 ± 10.53 | 40.11 ± 9.35 |
| STAI – Trait Anxiety | 32.21 ± 8.88 | 33.1 ± 9.44 | 31.22 ± 8.26 |

### Correlations Between HRV, Stress, and Burnout

Spearman correlation analysis demonstrated significant associations between HRV parameters and psychological distress (Figure 1). Lower RMSSD was correlated with higher depression levels (ρ = −0.281, p = 0.034) and greater suicidal ideation (ρ = −0.297, p = 0.025). Increased LF/HF ratio, indicating sympathetic dominance, was associated with higher emotional exhaustion (ρ = 0.324, p = 0.014). The SD1/SD2 ratio, a measure of autonomic rigidity, showed an inverse correlation with indolence (ρ = −0.316, p = 0.017). A lower overall HRV amplitude (Poincaré Area) was linked to higher depersonalization (ρ = −0.281, p = 0.034), reinforcing the role of autonomic regulation in occupational burnout. Table 3 summarizes the significant correlations observed in the study.

**Table 3.** Correlations Between HRV Parameters and Psychological Distress/Burnout Measures. Spearman’s rank-order correlations between heart rate variability (HRV) parameters and psychological distress/burnout measures. The table presents the mean ± standard deviation (SD) for each variable, the Spearman’s correlation coefficient (ρ), and the p-value for statistical significance (p < 0.05). Negative ρ values indicate an inverse relationship, while positive values indicate a direct correlation.

| HRV Parameter | Burnout/Stress Variable | Mean ± SD (HRV) | Mean ± SD (Burnout) | Spearman's $\rho$ | P value |
| --- | --- | --- | --- | --- | --- |
| RMSSD | Depression | 229.77 ± 104.86 | 10.50 ± 2.80 | -0.281 | 0.034 |
| RMSSD | Suicidal Ideation | 229.77 ± 104.86 | 28.10 ± 5.20 | -0.297 | 0.025 |
| LF/HF | Emotional Exhaustion | 0.89 ± 1.24 | 23.02 ± 8.72 | 0.324 | 0.014 |
| SD1/SD2 | Indolence | 0.83 ± 0.23 | 11.32 ± 3.78 | -0.316 | 0.017 |
| Poincaré Area | Depersonalization | 15.30 ± 4.50 | 9.13 ± 4.98 | -0.281 | 0.034 |

**Figure 1.**
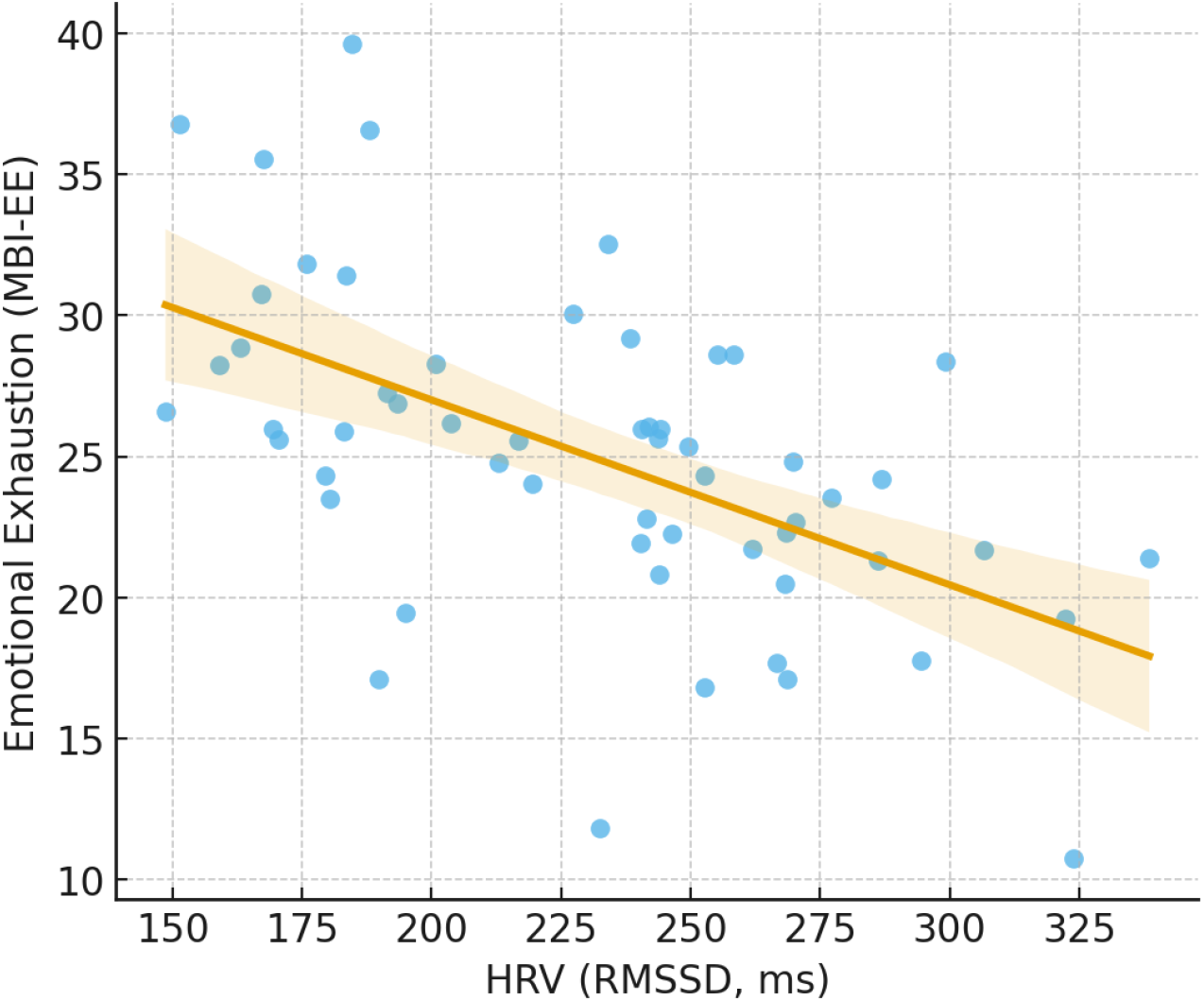
Correlation between HRV and Emotional Exhaustion. Scatterplot showing the negative correlation between HRV (RMSSD) and Emotional Exhaustion (MBI-EE). Lower HRV values are associated with higher levels of burnout, suggesting that autonomic dysfunction may be linked to occupational stress. The regression line represents the Spearman correlation trend.

### Workplace and Shift-Related Differences in HRV and Burnout

Comparisons between workplace settings and shift types revealed significant differences in burnout and autonomic regulation. ICU nurses exhibited lower HRV (RMSSD), and higher emotional exhaustion compared to general ward nurses (p < 0.05) (Figure 2). Similarly, night shift workers demonstrated significantly higher burnout levels and lower HRV values than day shift workers (p < 0.05) (Figure 3). When analyzing shift duration, individuals working 12-hour shifts exhibited better autonomic recovery, as indicated by higher PNN50 values (p = 0.044), compared to those on 8-hour shifts. These findings suggest that workplace conditions and shift schedules play a critical role in stress modulation. Table 4 presents the key group differences.

**Table 4.** Workplace and Shift-Related Differences in HRV and Burnout. Comparison of heart rate variability (HRV) parameters and burnout indicators across different work settings, shift types, and shift durations. The table presents the mean ± standard deviation (SD). “Higher in” indicates the group with the significantly greater value for the respective variable. p-values correspond to Mann-Whitney U or Kruskal-Wallis tests, with statistical significance set at p < 0.05.

| Comparison | Higher in | Mean ± SD<br>(Higher Group) | Mean ± SD<br>(Lower Group) | p-value |
| --- | --- | --- | --- | --- |
| RMSSD (ICU vs. Ward) | Ward | 210.50 ± 98.30 | 245.20 ± 110.70 | 0.037 |
| Emotional Exhaustion (ICU vs. Ward) | Ward | 25.80 ± 7.90 | 20.60 ± 8.40 | 0.041 |
| Burnout Index (Night vs. Day Shift) | Day Shift | 37.10 ± 4.20 | 33.40 ± 5.10 | 0.028 |
| HRV LF/HF Ratio (Night vs. Day Shift) | Day Shift | 1.02 ± 0.89 | 0.78 ± 0.65 | 0.039 |
| PNN50 (12h vs. 8h Shift) | 12h Shift | 62.30 ± 9.80 | 55.70 ± 11.20 | 0.044 |

**Figure 2.**
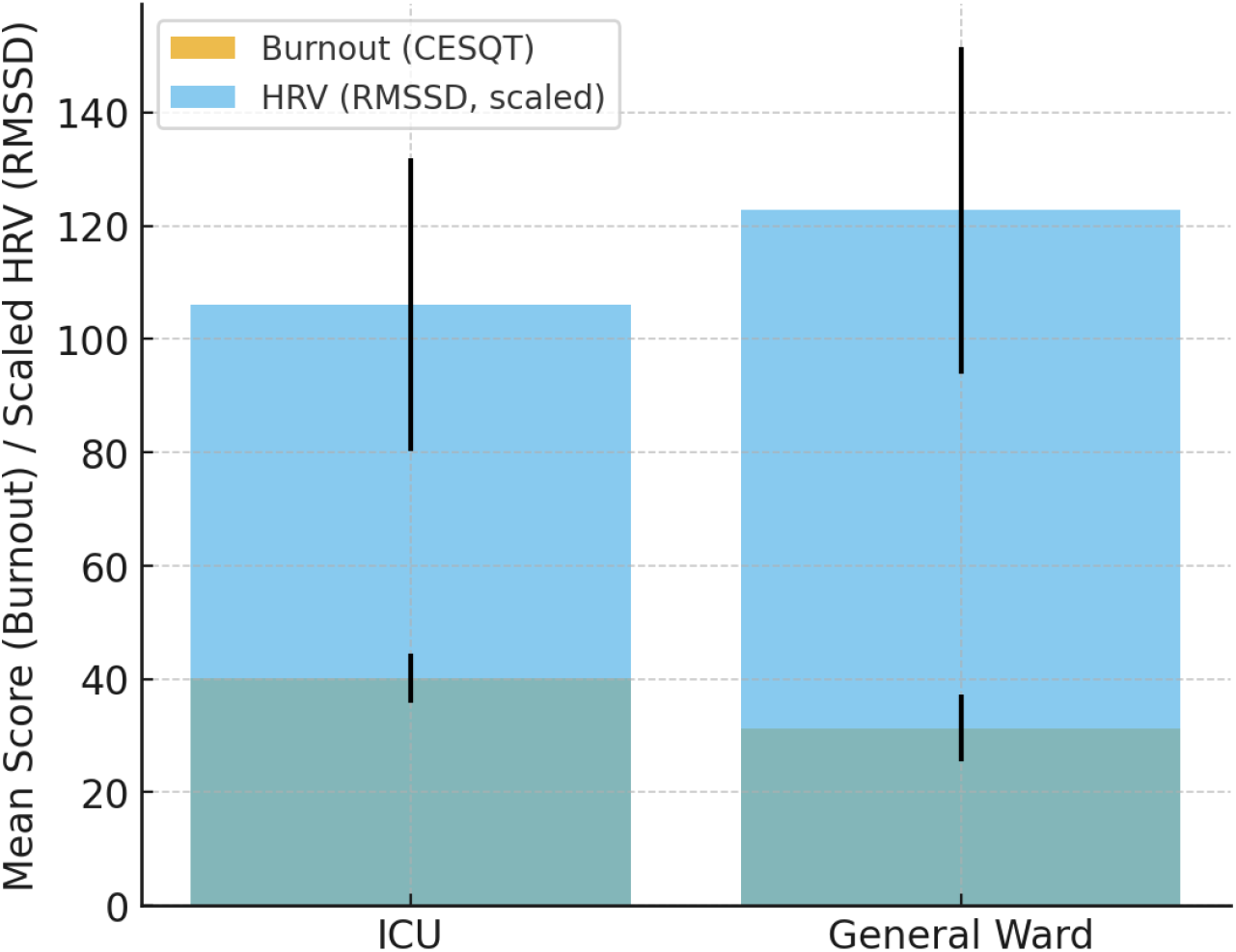
Burnout and HRV by Work Unit. Comparison of burnout (CESQT) and HRV (RMSSD) between ICU and general ward nurses. ICU workers exhibited higher burnout scores and lower HRV values, indicating a greater physiological stress burden. Error bars represent the standard deviation.

**Figure 3.**
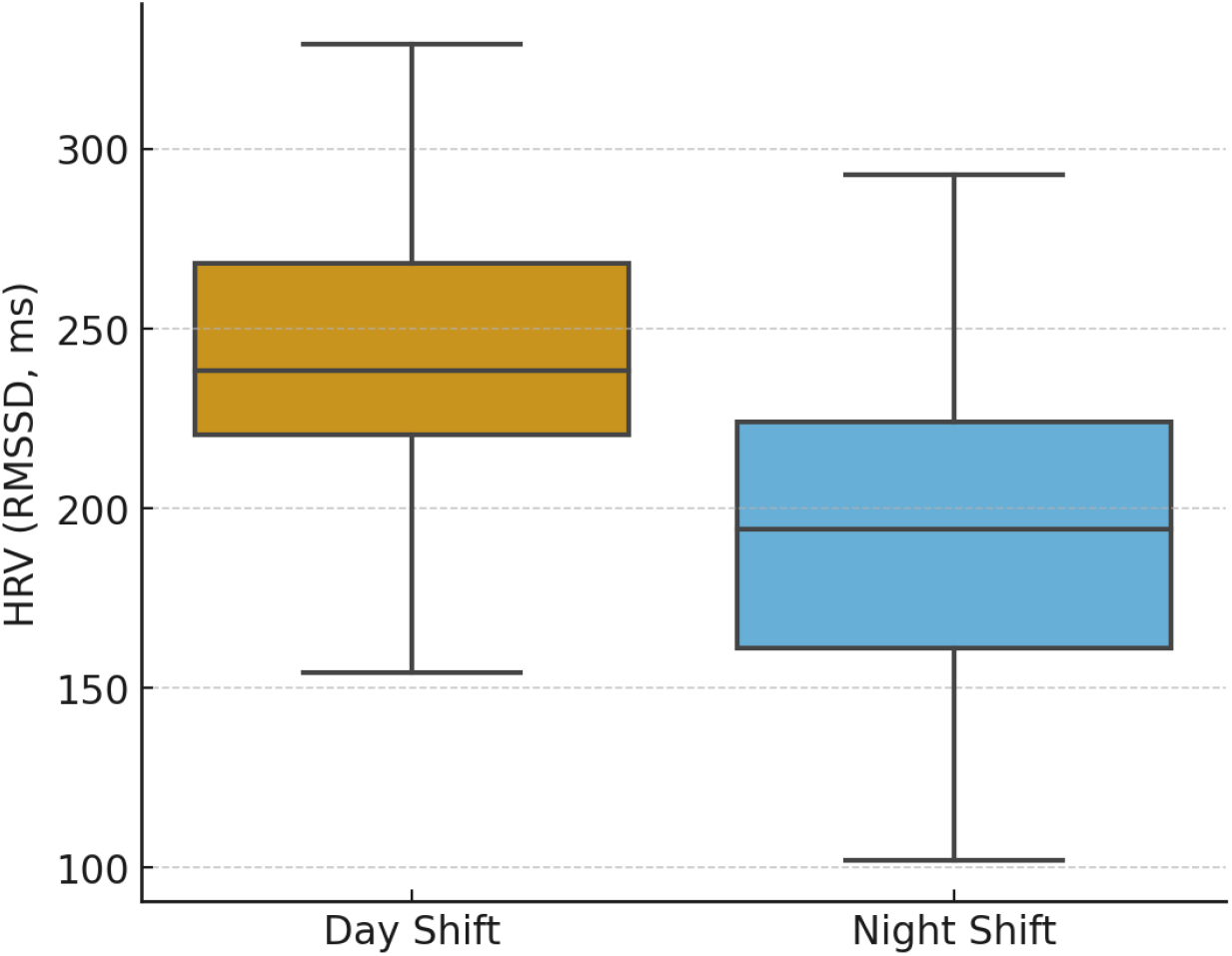
HRV Differences Between Day and Night Shift Workers. Comparison of heart rate variability (HRV) between day and night shift workers. Night shift workers exhibited lower HRV (RMSSD), indicating greater autonomic dysregulation and higher occupational stress compared to day shift workers. The boxplot represents the median and interquartile range (IQR), with whiskers extending to 1.5 times the IQR.

### Predictive Model for Burnout Risk

To assess the predictive value of HRV and psychological stress measures, two logistic regression models were developed. Model 1, which incorporated demographic and psychometric factors, achieved an AUC-ROC of 0.791 with an accuracy of 76.3%. Model 2, which integrated HRV parameters and occupational factors such as shift type and work unit, outperformed Model 1, reaching an AUC-ROC of 0.832 with an accuracy of 79.1% (Figure 4). The inclusion of objective HRV metrics significantly improved burnout risk assessment, reinforcing their role as potential physiological markers for occupational stress. Key predictors in Model 2 included the SD1/SD2 ratio, which emerged as the strongest biometric predictor of burnout (β = 0.37, p = 0.004), alongside occupational factors such as night shift work (*OR = 6.89, p = 0.027) and intention to leave the profession (*OR = 8.30, p < 0.001). Table 5 provides a comparative overview of both predictive models.

**Table 5.** Predictive Model Comparison. Performance metrics of two logistic regression models predicting burnout risk. Model 1 includes demographic and psychometric predictors, while Model 2 integrates HRV parameters and occupational factors. The table compares AUC-ROC, accuracy, sensitivity, and specificity expressed in percentages (%).

| Metric | Model 1 | Model 2 |
| --- | --- | --- |
| AUC-ROC | 0.791 | 0.832 |
| Accuracy (%) | 76.3 | 79.1 |
| Sensitivity (%) | 74.1 | 80.6 |
| Specificity (%) | 78.4 | 77.5 |

**Figure 4.**
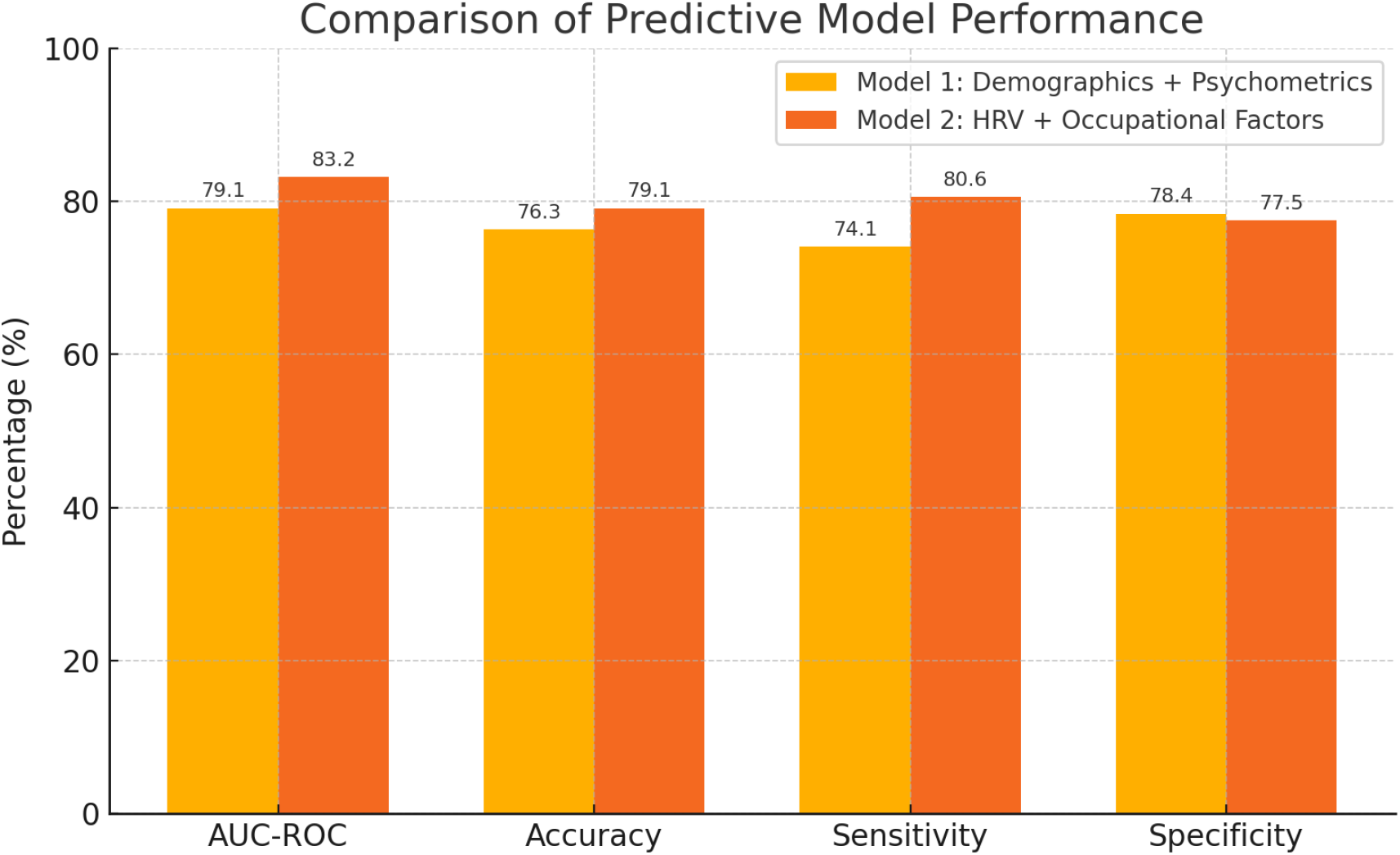
Comparative performance of two predictive models for burnout risk. Barplot showing the predictive performance of two logistic regression models based on 10-fold cross-validation. Model 1 includes demographic and psychometric variables (e.g., anxiety, burnout scores), while Model 2 incorporates heart rate variability (HRV) parameters and occupational factors (e.g., shift type). Model 2 outperforms Model 1 across all evaluated metrics, including AUC-ROC, accuracy, sensitivity, and specificity.

## DISCUSSION

This study evaluated heart rate variability (HRV) as an objective biomarker of burnout and developed predictive models to identify healthcare professionals at high risk of occupational stress. The findings reinforce the link between physiological stress responses, burnout symptoms, and work-related factors, providing a more objective framework for assessing occupational well-being [15].

The observed associations between HRV parameters—particularly SD1/SD2, RMSSD, and LF/HF—and burnout indicators support the hypothesis that chronic workplace stress is linked to autonomic nervous system dysregulation [16]. Specifically, lower SD1/SD2 and RMSSD values, reflecting reduced parasympathetic activity and impaired autonomic flexibility, were strongly correlated with higher emotional exhaustion, indolence, and depersonalization, consistent with the autonomic imbalance hypothesis, which posits that sustained workplace stress results in a shift towards sympathetic dominance and reduced vagal tone, thereby diminishing physiological resilience [17].

These findings are consistent with previous research demonstrating that healthcare professionals in high-demand environments, such as intensive care units and emergency departments, exhibit lower HRV compared to their counterparts in general wards. This autonomic imbalance is likely driven by chronic exposure to high-acuity patients, emotional distress, and disrupted sleep patterns, all of which contribute to heightened sympathetic activity and reduced stress recovery capacity [18]. Despite these well-documented associations, HRV has not yet been widely implemented as a screening tool for burnout in occupational settings, underscoring the need for further validation and integration into workplace health programs [19]. Previous research on burnout has primarily relied on self-reported psychometric assessments, particularly the Maslach Burnout Inventory (MBI). While the MBI remains widely used, its conceptual framework has been debated, particularly distinguishing depersonalization from emotional exhaustion. In contrast, this study employed the Cuestionario para la Evaluación del Síndrome de Quemarse por el Trabajo (CESQT), which includes additional dimensions such as indolence and guilt, two components that are particularly relevant in healthcare settings where moral distress and emotional detachment can be prevalent A key finding of this study was the significant variation in burnout and stress levels across different workplace conditions, reinforcing previous evidence on the negative impact of high-demand environments in healthcare [20]. Night shift workers and those with longer shifts (12 hours) exhibited lower HRV and higher burnout scores, indicating greater physiological and emotional strain. ICU nurses displayed more pronounced autonomic dysregulation compared to general ward nurses, possibly due to the intensity of critical care work, frequent exposure to trauma, and greater workload intensity. Additionally, participants who expressed an intention to leave the profession exhibited lower HRV values, reinforcing the strong relationship between job dissatisfaction and physiological stress responses.

These results align with prior research on shift work and burnout, which has demonstrated that circadian disruption, extended work hours, and high emotional demands contribute to increased allostatic load and reduced stress recovery capacity [21].

The present findings are consistent with previous studies that have established HRV as a physiological correlation of psychological distress [22]. Lower SD1/SD2 ratios and reduced HRV amplitude have been widely observed in individuals experiencing occupational burnout, depression, and chronic stress-related disorders. The observed positive correlation between LF/HF ratio and emotional exhaustion also aligns with research demonstrating that sympathetic overactivation contributes to burnout-related fatigue.

Regarding shift work, the observed lower HRV in night shift workers corroborate earlier findings linking circadian rhythm misalignment to autonomic dysfunction. Disruptions in melatonin secretion, cortisol regulation, and cardiovascular function have been documented in night shift personnel, increasing the risk of stress-related disorders. These physiological disruptions may explain why night shift workers exhibit a higher prevalence of burnout symptoms and poorer mental health outcomes [23].

A key contribution of this study is the development of a predictive model for burnout risk, integrating HRV and occupational factors [24]. The superior performance of Model 2 (AUC-ROC = 0.832), which incorporated HRV parameters, shift type, and work-related stressors, demonstrates that objective physiological markers enhance burnout prediction compared to traditional demographic and psychometric models. These findings suggest that HRV monitoring could be incorporated into occupational health programs as a non-invasive, objective screening tool for early burnout detection. Wearable HRV devices, for example, could enable continuous physiological monitoring, allowing for real-time interventions before burnout symptoms escalate. By eliminating subjective bias from self-reported stress assessments, HRV-based screening could provide a more reliable and reproducible method for identifying high-risk individuals.

### Implications for Occupational Health and Clinical Practice

The identification of HRV as a physiological marker of burnout has important clinical and occupational health implication [25]. The integration of real-time biometric monitoring in healthcare settings —such as wearable HRV devices—could facilitate early detection of autonomic dysregulation, enabling timely interventions before burnout progresses to more severe psychological or physical conditions [26]. Objective physiological screening could complement traditional psychometric evaluations by reducing subjectivity and enhancing reproducibility.

Given the higher stress burden observed in ICU professionals and night shift workers, targeted interventions should focus on optimizing staffing patterns, revising shift schedules, and implementing structured recovery protocols to mitigate the physiological impact of chronic workplace stress [27]. Strategies such as limiting consecutive night shifts, ensuring adequate rest periods, and reducing excessive workloads may help preserve autonomic function and improve overall well-being [28]. Additionally, stress reduction programs, including mindfulness-based interventions, resilience training, and HRV biofeedback techniques, could enhance autonomic flexibility and reduce burnout symptoms [29].

At an institutional level, these findings underscore the need for policies that integrate objective physiological assessments into occupational health frameworks [30]. Traditional burnout assessments, which rely heavily on self-reported measures, may benefit from the incorporation of HRV-based monitoring tools to provide a more data-driven and objective approach to workforce well-being. Routine HRV screenings in occupational health evaluations could facilitate the early identification of high-risk individuals, ultimately improving staff retention, job satisfaction, and patient care quality.

### Limitations and Future Directions

This study presents several limitations that should be acknowledged. First, the relatively small and heterogeneous sample (N = 57) may limit the generalizability of findings and restrict the statistical power to explore interactions between variables such as age, gender, job role, or tenure. Although exploratory comparisons were conducted, larger samples are needed to enable multivariable adjustment and subgroup analyses.

Second, the cross-sectional design limits causal inference regarding the relationship between HRV and burnout. Longitudinal studies are needed to clarify the temporal dynamics of autonomic dysregulation in occupational stress.

Third, HRV parameters were derived from short-term recordings under resting conditions. While time-domain and frequency-domain indices (e.g., RMSSD, LF/HF) are valid in this setting, nonlinear markers such as the SD1/SD2 ratio should be interpreted cautiously and viewed as exploratory.

Fourth, certain contextual factors—such as sleep quality, cumulative workload, and coping strategies—were not fully assessed or included in predictive models due to sample size constraints. These variables should be considered in future research.

Finally, although the predictive model showed promising accuracy using internal cross-validation, its generalizability remains to be confirmed through external validation in larger and more diverse cohorts. Future work should also evaluate the feasibility of implementing HRV-based monitoring tools for early identification and intervention in occupational health programs.

## CONCLUSION

This study highlights the potential utility of heart rate variability as a physiological marker for identifying healthcare professionals at risk of burnout. HRV-based predictive models outperformed those relying solely on demographic and psychometric data, suggesting that the integration of objective physiological data can enhance early detection strategies for occupational stress.

The findings support the incorporation of HRV monitoring into occupational health frameworks as a complementary tool to traditional self-report measures. Real-time biometric assessment may facilitate more timely and personalized interventions, contributing to improved well-being, reduced turnover, and sustained quality of care in high-demand healthcare environments.

Future research should prioritize external validation of these predictive models in larger, multicenter samples and evaluate the feasibility, acceptability, and impact of HRV-based screening tools in routine clinical practice.

## List of abbreviations

AIC: Akaike Information Criterion
AUC-ROC: Area Under the Receiver Operating Characteristic Curve
BIC: Bayesian Information Criterion
Brier Score: Metric to evaluate model calibration
CESQT: Cuestionario para la Evaluación del Síndrome de Quemarse por el Trabajo
CESQT Burnout Index: Composite measure of burnout (aggregated CESQT score)
CESQT-CU: Guilt (Culpa)
CESQT-DP: Psychological Exhaustion (Desgaste Psíquico)
CESQT-IN: Indolence (Indolencia)
CESQT-IT: Work Engagement (Implicación en el Trabajo)
DL: Dyslipidemia
DM: Diabetes Mellitus
HRV: Heart Rate Variability
HTN: Hypertension
ICU: Intensive Care Unit
LF/HF: Low-Frequency to High-Frequency Ratio (indicator of autonomic balance)
MANOVA: Multivariate Analysis of Variance
MBI: Maslach Burnout Inventory
MBI-AE: Emotional Exhaustion (Agotamiento Emocional)
MBI-DP: Depersonalization (Despersonalización)
MBI-RP: Personal Accomplishment (Realización Personal)
MSE: Mean Squared Error
PNN20: Percentage of adjacent NN intervals differing by more than 20 ms
PNN50: Percentage of adjacent NN intervals differing by more than 50 ms
ProQOL: Professional Quality of Life Scale
ProQOL-AM: General Burnout (Burnout General)
ProQOL-AP: Burnout Fatigue (Agotamiento Profesional)
ProQOL-ETS: Secondary Traumatic Stress (Estrés Traumático Secundario)
ProQOL-SC: Compassion Satisfaction (Satisfacción por Compasión)
rMSSD: Root Mean Square of Successive Differences (HRV parameter)
SD1/SD2: Ratio of short-term (SD1) to long-term (SD2) variability in HRV
STAI: State-Trait Anxiety Inventory
STAI-AE: State Anxiety (Ansiedad Estado)
STAI-AR: Trait Anxiety (Ansiedad Rasgo)

## Acknowledgments

We sincerely thank all the nurses and healthcare assistants who participated in this study. Your contributions were invaluable, and this research would not have been possible without your involvement. We appreciate your time and effort in helping us better understand the stress and burnout experienced in ICU settings

## Authors’ Contributions

ARL designed the study, performed the statistical analysis, and wrote the manuscript as the principal investigator; TSP collected the data; RGC and ARN participated in the study design and statistical analysis.

## Data Availability

To support reproducibility and facilitate further research, we have made the following resources available: **Biometric Data:** The anonymized biometric data collected during the study, including heart rate variability (HRV) parameters (e.g., rMSSD, LF/HF ratio), is available through Fighshare at https://doi.org/10.6084/m9.figshare.28378298.v1

## Supplementary Material Appendix A

Complete versions of the following validated instruments used in the study:

- Cuestionario para la Evaluación del Síndrome de Quemarse por el Trabajo (CESQT)
- Maslach Burnout Inventory (MBI)
- Professional Quality of Life Scale (ProQOL)
- State-Trait Anxiety Inventory (STAI)

